# Heat-related mortality attributable to climate change across age and sex in the Netherlands, 1971–2019

**DOI:** 10.64898/2026.09.03.26362192

**Authors:** Adam Finnemann, Fabian Dablander, Reinout Wiers, Jonas Haslbeck

## Abstract

Heat-related mortality is increasingly attributed to human-caused climate change, but most estimates are aggregated across time and population groups, masking who carries the burden and how that distribution shifts. We estimated heat-related deaths attributable to climate change in the Netherlands from 1971 to 2019 disaggregated by age and sex using a model in which the temperature–mortality relationship changes continuously from year to year. Among adults aged 65 and over, heat caused an average of 644 deaths per warm season, of which 381 (59%) would not have occurred without global warming. This fraction rose from 46% in 1971–1994 to 79% in 2010–2019. The burden falls overwhelmingly on the oldest: women aged 80 and over account for 55% of climate-change-attributable deaths and men of the same age for 24%, leaving 21% for the 65–79 groups. Per-capita rates at these ages are now similar in both sexes, so the female burden reflects women outliving men rather than greater susceptibility. Over the study period the standardized heat-mortality rate fell by more than half, because vulnerability declined faster than exposure rose. That decline began decades before the 2007 Dutch Heat Plan, which argues against the plan as the main driver, and it followed different paths in men and women. The decline in heat vulnerability must be better understood if it is to be sustained and extended against the unprecedented heat ahead. At the same time, we must urgently limit climate change to prevent further loss of life.

## 1 Introduction

Human-caused climate change has raised global mean surface temperature by about 1.4 ℃ above pre-industrial levels, with disastrous impacts (Forster et al., 2026). The field of attribution science quantifies how much of a given impact is due to global warming (Mitchell et al., 2016; Otto, 2023; Stott et al., 2004). It has shown that the record-breaking European heatwaves of summer 2026 were greatly amplified by climate change (World Weather Attribution, 2026). In Europe, they have caused widespread wildfire destruction and an estimated 10,000 excess deaths (Euronews, 2026; World Meteorological Organization, 2026; World Weather Attribution, 2026). In the United States, a succession of heat domes has driven temperatures to record-challenging highs, killing at least 70 people in July (CNN, 2026; Kaplan, 2026). Across 43 countries, about 37% of warm-season heat deaths are already attributable to global warming (Vicedo-Cabrera et al., 2021), a conclusion echoed across the subsequent literature (Beck et al., 2024; Hundessa et al., 2025; Stuart-Smith et al., 2025; Wan et al., 2025). In Germany, this share has been rising by roughly 5.6 percentage points per decade (Huber et al., 2025).

Quantifying the impacts of global warming is important, for example to raise public awareness and to provide evidence for climate litigation (Stuart-Smith et al., 2021). Attribution findings are increasingly featured in media coverage of extreme weather (Hopke & Wozniak, 2025; Osaka et al., 2020), and public attribution of such events to climate change is associated with greater support for climate policy (Cologna et al., 2025). Climate litigation holds states and companies accountable for climate impacts (Setzer & Higham, 2025). For instance, elderly Swiss women, organized in *KlimaSeniorinnen*, won a case before the European Court of Human Rights by showing that weak climate policy exposed them to increased heat-related mortality (Savaresi, 2025).

Litigation claims often depend on showing that a particular group bears a burden, yet most attribution evidence is aggregate (Vicedo-Cabrera et al., 2021). Heat risk falls disproportionately on the elderly and on women (Achebak et al., 2019; Folkerts et al., 2021), rising steeply with age as thermoregulation declines and other health conditions accumulate (Bunker et al., 2016; Ebi et al., 2021). In the European summer of 2022, adults aged 80 and over accounted for 60% of heat deaths, and heat-related mortality was 56% higher among women than men (Ballester et al., 2023). Long-term attribution of heat mortality to climate change, however, has never been analyzed by age and sex. Only single events, such as the 2022 European summer, have been analyzed in this way (Beck et al., 2024; Vicedo-Cabrera et al., 2023).

Huber et al. (2025) provide the first long-term, time-varying attribution of heat mortality, showing that the share of heat deaths attributable to climate change has grown over recent decades. Time-varying methods matter because both exposure and population vulnerability changed over this period (Gasparrini, Guo, Hashizume, Kinney, et al., 2015; Vicedo-Cabrera et al., 2018). They also allow the study of dynamics around critical events and policy interventions such as heat plans, which helps evaluate their effectiveness. We focus on the Netherlands, where the 2007 National Heat Plan alerts healthcare systems, interest organizations, and the public when heatwaves are forecast (Klompmaker & Hagens, 2025; Kunst & Britstra, 2013). Similar policies now operate in many countries, and whether they meaningfully reduce heat deaths remains unclear (Boeckmann & Rohn, 2014; Urban et al., 2025). Here, we build on the work of Huber et al. (2025) in two ways. We resolve deaths by age and sex, and we use a fully continuous model across the study period (1971–2019). This allows us to establish how much of Dutch heat mortality is attributable to climate change, which age and sex groups bear that burden, and how heat vulnerability evolved across five decades spanning the introduction of the National Heat Plan.

## 2 Methods

We estimated heat-related mortality attributable to human-caused climate change in the Netherlands, stratified by age groups (0–64, 65–79, 80+) and sex, over the period 1971–2019. We define climate-change-attributable deaths as the difference between the heat-attributable deaths under observed temperatures and those under a counterfactual temperature series representing a world without global warming. The analysis follows recent observation-based health-attribution methods (Stuart-Smith et al., 2025; Vicedo-Cabrera et al., 2021) and proceeds in three stages, which we outline here and describe more fully below. Appendix A gives the full technical specification of each stage, covering the data sources, the model settings, and the propagation of uncertainty.

**Stage 1: Epidemiological analysis.** In this stage, we estimate how heat affects mortality (purple in Figure 1). For each age *×* sex stratum, we fit a single distributed lag non-linear model (DLNM) linking weekly warm-season temperature to weekly all-cause mortality. This yields year- and stratum-specific exposure–response curves describing how mortality changes with temperature in each year.

**Figure 1:**
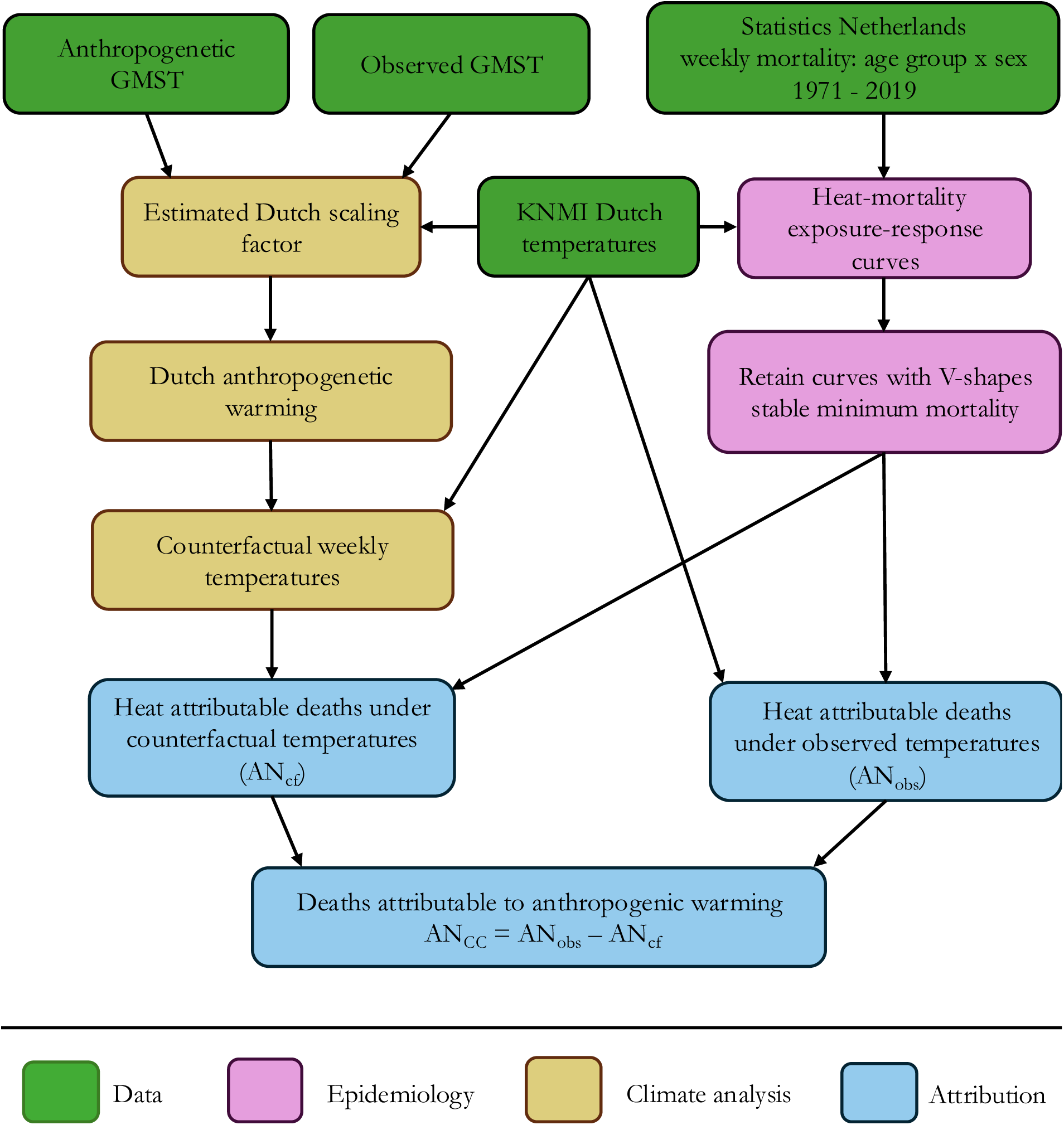
Overview of the three-stage analytical framework. Stage 1 (purple): epidemiological modeling of the temperature–mortality association. Stage 2 (yellow): construction of counterfactual temperatures without global warming. Stage 3 (blue): attribution of heat-related deaths to climate change. The main data sources are listed in green (GMST: global mean surface temperature; KNMI: Royal Netherlands Meteorological Institute).

**Stage 2: Constructing counterfactual temperatures.** Here, we construct a counterfactual temperature series representing Dutch temperatures without global warming (yellow in Figure 1). We base this on published estimates of global warming at the global level, which we scale to the Netherlands using a scaling factor estimated from monthly temperature data. Subtracting the resulting Dutch warming from the observed weekly series yields a counterfactual that removes the anthropogenic signal while preserving observed week-to-week variability.

**Stage 3: Attribution of heat-related mortality to climate change.** In this stage, we combine each stratum’s year-specific exposure–response curves with the observed and counterfactual temperatures to estimate how many deaths heat caused under each scenario (blue in Figure 1). The difference between the two is our estimate of the deaths attributable to climate change. Uncertainty from the exposure–response model and from the counterfactual is carried through jointly using Monte Carlo simulation. We describe each stage in more detail below.

All analyses were conducted in R version 4.5.3 (R Core Team, 2024). Distributed lag non-linear models were fit with mgcv (v1.9-4) (Wood, 2017) and dlnm (v2.4.10) (Gasparrini, 2011). Monte Carlo draws from the coefficient covariance matrix used MASS (v7.3-65) (Venables & Ripley, 2002). Data preparation used dplyr (v1.2.1) (Wickham et al., 2023) and figures were produced with ggplot2 (v4.0.2) (Wickham, 2016).

### 2.1 Data sources

Weekly all-cause mortality counts by age group and sex, and annual population counts used as an offset, were obtained from Statistics Netherlands (CBS). Weekly mean and maximum temperatures were constructed from national station data provided by the Royal Netherlands Meteorological Institute (KNMI). The anthropogenic warming component comes from the Indicators of Global Climate Change (IGCC) project (Forster et al., 2025), and monthly indices of the El Niño–Southern Oscillation and the North Atlantic Oscillation from NOAA. All data are freely and publicly available, with archived versions and download scripts provided in our OSF repository (https://osf.io/d6rme). Dataset identifiers and access details are listed in Appendix A.1.

### 2.2 Stage 1: Epidemiological analysis

#### Study design and stratification

We use weekly death counts across the warm season in the Netherlands, separately for men and women in three age groups, giving six strata in total. The analysis is restricted to the warm season (May–September) to focus on heat-related death. We end the series in 2019 to avoid COVID-related confounds in mortality rates from 2020 onward. The single warm-season week containing the 2014 MH17 air disaster, an external mortality shock unrelated to temperature, was replaced by the average of the four surrounding weeks.

#### Exposure definition

Heat exposure is represented by weekly mean temperature, *T*_mean_(*t*), obtained by averaging daily mean temperatures from KNMI national station data within each ISO week. This variable is the primary exposure in all main analyses. As a sensitivity analysis, we repeat the modeling with weekly maximum temperature, *T*_max_(*t*), defined as the highest daily maximum within the week (see Appendix B.1).

#### Time-series regression model

We fit one quasi-Poisson generalized additive model per stratum over the full period 1971–2019, with a population offset, using a quasi-Poisson family to account for the over-dispersion typical of weekly death counts. This follows the DLNM framework of Gasparrini and Leone (2014), used widely in temperature-mortality assessments (Gasparrini, Guo, Hashizume, Lavigne, et al., 2015; Vicedo-Cabrera et al., 2021) and climate-change health attribution (Stuart-Smith et al., 2025). Weekly deaths depend on four quantities: population size, recent temperature, time of year, and a slow-moving trend capturing gradual change in mortality rates unrelated to heat, such as improvements in medical care.

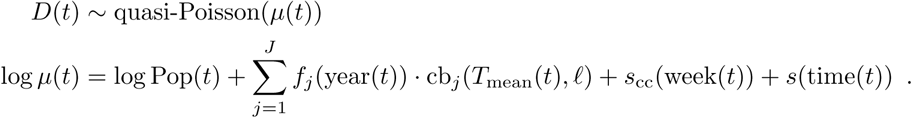

*D*(*t*) is the weekly death count and log Pop(*t*) the log population offset. The cb*_j_*(*·*) are the *J* columns of a quadratic B-spline basis over temperature and lags *ℓ* = 0, 1, 2 weeks, which together let temperature act non-linearly and with delay. Each column is multiplied by its own smooth function of calendar year, *f_j_*(year(*t*)). This is what lets the whole exposure–response curve change shape over time, including the minimum-mortality temperature and the slope at high temperatures. The remaining two terms control for time. *s*_cc_(week(*t*)) is a cyclic cubic spline of week-of-year, absorbing seasonality within the warm season, and *s*(time(*t*)) a smooth of calendar time, absorbing slow change in baseline mortality. The choice of lag window and the flexibility of both time terms are set out in Appendix A.2, and varied as a robustness check in Appendix B.2.

### 2.3 Stage 2: Constructing counterfactual temperatures

The anthropogenic warming component is taken from the Global Warming Index (Haustein et al., 2017), updated through the Indicators of Global Climate Change project (Forster et al., 2025), and linked to Dutch temperatures via a season-specific GMST scaling factor. This approach originates in probabilistic event-attribution frameworks (Philip et al., 2020; van Oldenborgh et al., 2021) and has recently been applied to health attribution (Stuart-Smith et al., 2025).

#### Global warming signal

We obtain annual estimates of anthropogenic global mean surface temperature change from the IGCC/GWI time series, which splits total warming in year *y* into a natural and an anthropogenic component, ΔGMST_NAT_(*y*) and ΔGMST_ANT_(*y*) (Forster et al., 2025):

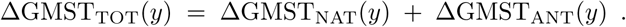

We define ΔGMST_ANT_(*y*) as the sum of the contributions from greenhouse gases and from other human influences on the climate. This is the signal to be removed from observed Dutch temperatures, after translating it to the Netherlands scale as described below. Its smoothing and percentile treatment follow standard attribution protocol (Philip et al., 2020; van Oldenborgh et al., 2021) and are detailed in Appendix A.3.

#### Translating global warming to the Netherlands: the scaling factor *α*

Global warming does not raise temperatures everywhere at the same rate, so we need to know how much Dutch temperatures rise for each degree of global warming. We call this the scaling factor *α* and estimate it by regressing monthly Dutch mean temperature on the annual global warming level ΔGMST_ANT_, controlling for El Niño–Southern Oscillation (Niño3.4) and the North Atlantic Oscillation, absorbing the within-season seasonal cycle with month fixed effects, and clustering standard errors by year:

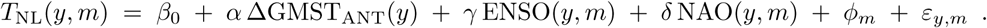

Here *y* and *m* index year and calendar month, *ϕ_m_*is a month fixed effect, and *ε_y,m_* the residual. The regression is fitted on warm-season months (May–September), and *α* is the coefficient of interest, the degrees of Dutch warm-season warming per degree of global warming. Dutch anthropogenic warming in year *y* is then Δ*T*_NL,ANT_(*y*) = *α ·* ΔGMST_ANT_(*y*), and counterfactual weekly temperatures are obtained by subtracting this shift from the observed series. For week *t* falling in year *y*(*t*),

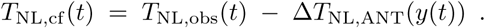

By construction, *T*_NL,cf_ (*t*) retains the observed week-to-week variability and event timing, but represents warm-season temperatures in a world without global warming (Stuart-Smith et al., 2025). The constant-*α* assumption is examined in dedicated robustness checks (Appendix B.3).

#### Uncertainty propagation

Uncertainty in the counterfactual temperatures has two sources, the anthropogenic GMST estimate and the scaling factor *α*. We combine them by Monte Carlo simulation and pass the mean and standard deviation of the resulting annual shift Δ*T*_NL,ANT_(*y*) forward as the climate-uncertainty input to the attribution stage (Section 2.4; details in Appendix A.3).

### 2.4 Stage 3: Attribution of heat-related mortality to climate change

For every year and each sex *×* age-group stratum, we compute the heat-attributable number of deaths (AN_heat_) in the observed world and the corresponding number in the counterfactual world without global warming. Heat-attributable deaths are the excess deaths above the minimum mortality temperature (MMT), summed over lags of 0–2 weeks, with the MMT taken as the year-specific risk-minimizing temperature, consistent with standard practice in heat-attribution studies (Gasparrini, Guo, Hashizume, Lavigne, et al., 2015; Stuart-Smith et al., 2025; Vicedo-Cabrera et al., 2021). Writing AN_heat,cf_ for the corresponding quantity under counterfactual temperatures, climate-change-attributable deaths are

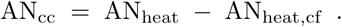

We report national totals as age–sex standardized rates per 100,000. Raw national counts rise over time simply because the population at risk grew and aged. To obtain a measure comparable across years, we divide each stratum’s attributable deaths by its own population and combine the resulting rates by direct standardization, holding the sex *×* age structure fixed at its 1971–2019 average. We also report the annual climate-change share of heat-attributable deaths (AN_cc_*/*AN_heat_) and summarize its trend (Appendix B.6).

#### Joint uncertainty propagation

Uncertainty in the exposure–response function and in the counterfactual temperatures is propagated jointly via Monte Carlo. Within each draw, AN_heat_ and AN_heat,cf_ are computed from the *same* coefficient draw, so that AN_cc_ is a paired difference. Intervals are the 5th and 95th percentiles of the draws, with the 50th as the central estimate, and national intervals are formed by summing stratum-specific draws before taking percentiles (Appendix A.4).

#### Null-mortality robustness

A well-behaved method should return close to zero attributable deaths when mortality is in fact unrelated to temperature. Applying the method to a synthetic series in which weekly deaths were independent of temperature showed that it is unstable under this null, with the MMT frequently snapping to boundary values (Appendix B.4). We therefore report and inspect the fitted relative-risk curves and MMT of all strata to confirm that each was well defined (Appendix B.7).

## 3 Results

We report the counterfactual temperature estimates in Section 3.1, the overall heat and climate-change-attributable mortality in Section 3.2, the heat and climate-change-attributable mortality by sex and age in Section 3.3, and robustness checks of central assumptions in Section 3.4.

### 3.1 Counterfactual temperatures

By 2019, observed Dutch warm-season temperatures exceeded the counterfactual without anthropogenic warming by approximately 2.5 ℃ (Figure 2).

**Figure 2:**
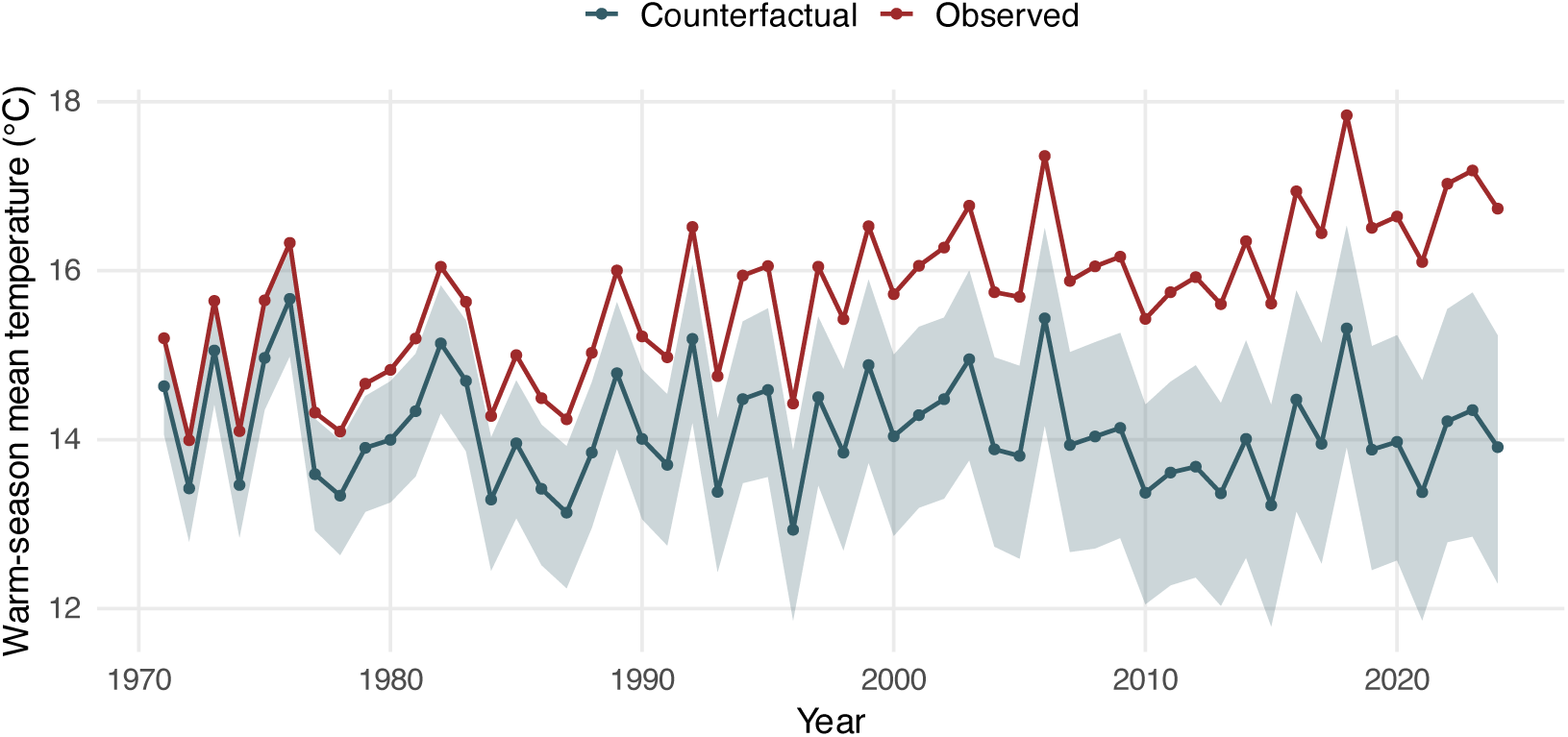
Observed (red) versus counterfactual (blue) warm-season mean temperature, 1971–2019. The widening gap reflects the growing global warming signal. Shaded band shows the 90% uncertainty interval around the counterfactual.

### 3.2 National heat and climate-change-attributable mortality

Our main strata are age groups 65 or older, because the exposure–response curves below age 65 were near-linear with unstable, boundary-snapping minimum mortality temperatures, precluding reliable estimation in that group (see Appendix B.7). Over the 49 warm seasons from 1971 to 2019, 1,995,150 all-cause deaths were recorded in the main strata of this study, with an average of 40,717 deaths per warm season. The average death count rose by roughly 44% over the study period, from 32,805 deaths per warm season in 1971–1980 to 47,164 in 2010–2019. This increase tracks demographic change. The population in these age groups grew roughly 2.5-fold over the same period, from 1.34 million people in 1971 to 3.34 million in 2019.

Across 1971–2019, heat was responsible for an average of 644 deaths per warm season, of which 381 (59%) were attributable to global warming. Over the whole period this amounts to 31,600 heat-attributable and 18,700 climate-change-attributable deaths. The climate-change share rose steadily across the three periods (1971–1994: 46%; 1995–2009: 66%; 2010–2019: 79%; Appendix B.6), while total heat-attributable deaths peaked in 1995–2009 (862/yr) before falling to 483/yr in 2010–2019, after averaging 575/yr in 1971–1994.

The population in these age groups grew and aged over the study period, so absolute counts conflate demographic change with changes in exposure and vulnerability. We therefore express the burden as a standardized rate with the sex *×* age structure held fixed at its study-period average (reported in Figure 3). On this basis the heat-mortality rate fluctuated around 37 per 100,000 between 1971 and 2009 (decade means 27 to 47), before dropping to 15 per 100,000 in 2010–2019. The climate-change-attributable rate followed a different path, roughly doubling from 13 per 100,000 in the 1970s to a peak of 27 in the 1990s before falling to 12 per 100,000 in 2010–2019.

**Figure 3:**
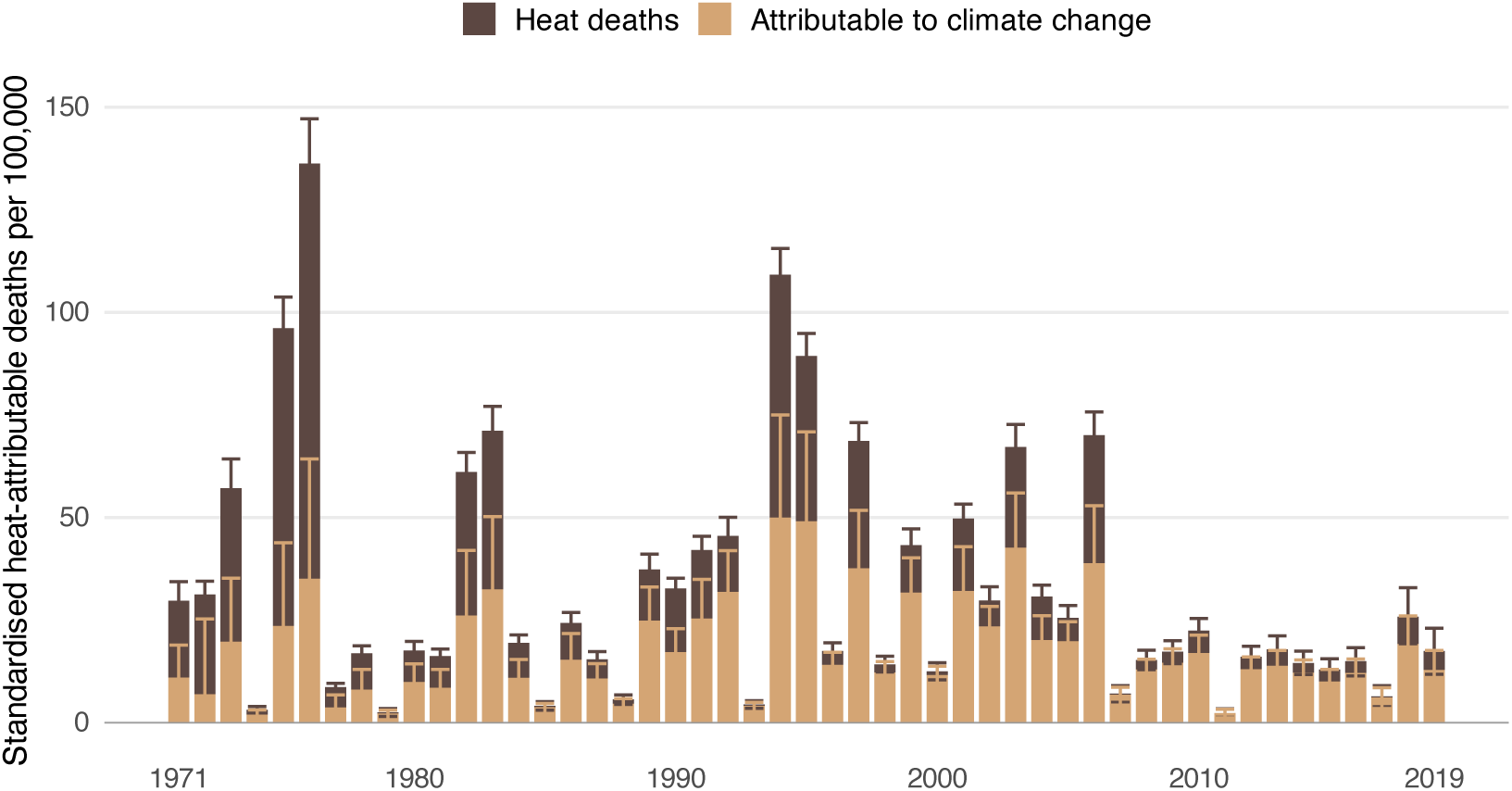
Standardized heat-attributable deaths per 100,000 per warm season in the Netherlands (1971– 2019), decomposed into deaths attributable to climate change (sand) and remaining heat deaths (dark brown). Rates are age-standardized to the period-average age–sex structure. Error bars show 90% uncertainty intervals.

### 3.3 Who bears the burden: age and sex breakdown

Heat-attributable mortality is concentrated in adults aged 65 or older, and it has declined over time in each of these strata, although among women aged 80+ the decline set in only after a rise that peaked around 2000. In absolute counts, annual heat deaths halved among women aged 80+ between 1995–2009 and 2010–2019, from 538 to 266 per year, and fell more modestly among men aged 80+, from 189 to 140 per year. Women aged 65–79 declined from 81 per year in 1971–1994 to 20 per year in 2010–2019.

These counts reflect population size as well as risk. On a per-capita basis (Figure 4), the burden falls overwhelmingly on the oldest adults, with heat-mortality rates at 80+ roughly ten times those at 65–79. Averaged over the study period, rates within the 80+ group are similar for men and women, even though absolute deaths are much higher among women.

**Figure 4:**
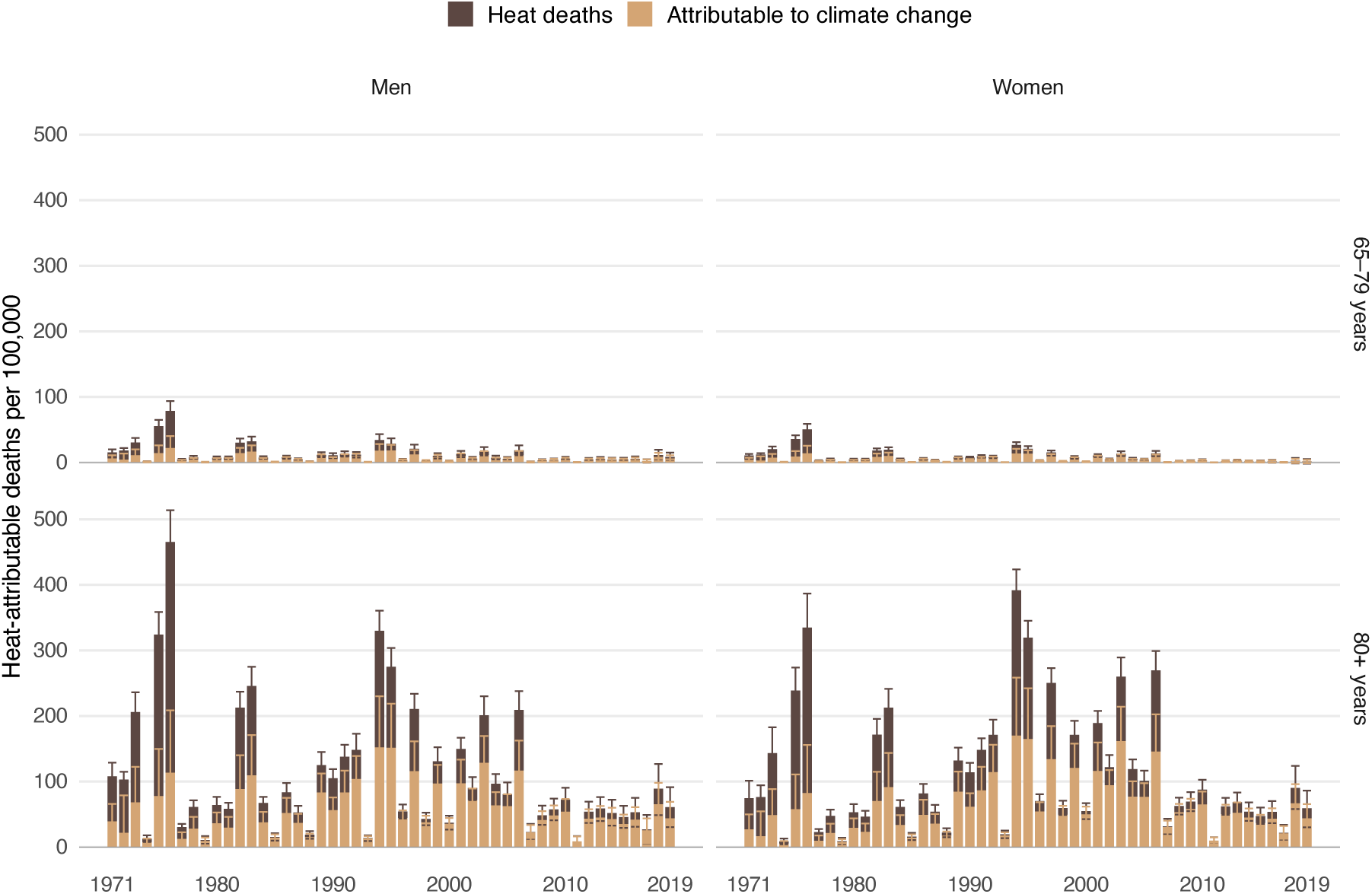
Heat-attributable deaths per 100,000 per warm season by sex and age group (1971–2019), decomposed into deaths attributable to climate change (sand) and remaining heat deaths (dark brown). Rates are per 100,000 of each stratum’s own population. Error bars show 90% uncertainty intervals. Only strata with a robust temperature–mortality association are shown.

The climate-change share has risen in every stratum. Over the study period, women aged 80+ account for 55% of all climate-change-attributable deaths (cumulative *∼*10,200) and men aged 80+ for a further 24% (*∼*4,500); the two 65–79 groups make up the remaining 21% (men 12%, *∼*2,200; women 9%, *∼*1,700).

Heat vulnerability at ages 80 or older followed different trajectories in men and women. The male heat-mortality rate fell steadily throughout, from 145 per 100,000 in 1975 to 53 in 2019, a decline of 63%. The female rate instead rose from 97 per 100,000 in 1975 to a peak of 150 in 2000, before falling by 71% to 44 in 2019. The sexes therefore differed most around 2000, when female rates exceeded male rates by 44%, but the subsequent decline in women has brought the two to comparable levels. No such crossover occurs at ages 65–79, where female rates remained below male rates throughout and the gap widened. Rates for selected years are given in Appendix Table S2.

### 3.4 Robustness

To assess whether our central findings depend on specific modeling choices, we test them against two alternative specifications. We replace weekly mean temperature with weekly maximum temperature (Appendix B.1), and we vary the controls for the seasonal cycle (Appendix B.2). Under both alternatives, our central results of a rising climate-change share, a declining total heat burden, and a concentration of that burden among women aged 80+ are robust. The fitted time-varying relative-risk curves and minimum mortality temperatures underlying these results, for every stratum, are reported in Appendix B.7.

Because the scaling factor *α* sets the magnitude of the counterfactual cooling, it is the single most influential input to the attribution, and we examine it in two ways. First, we test whether *α* is constant across time. Tests of its stationarity over time and across the temperature distribution show no meaningful variation (Appendix B.3). Second, we assess how strongly the results depend on its value by re-running the attribution across the 95% confidence interval of *α* (Appendix B.5). The magnitude of the attributed share scales with *α*, but in every case the share stays positive, rises monotonically across the three periods, and preserves the age–sex ordering, with women aged 80+ accounting for 55% of climate-change-attributable deaths and men aged 80+ for 24%.

## 4 Discussion

This paper provides the first long-term attribution of heat-related mortality to climate change resolved by age and sex, covering 1971–2019. Our central finding is that climate change accounts for a growing share of a shrinking burden. The climate-change share rose from 46% to 79% over the study period, while the standardized heat-mortality rate fell from 37 to 15 per 100,000. The majority of heat deaths in the Netherlands are therefore due to human activity, primarily the burning of fossil fuels (Forster et al., 2025). Emissions must consequently fall to limit future heat deaths, and those who continue to emit can be held accountable for the harm already caused (Setzer & Higham, 2025).

Vulnerability to heat fell in every age group. While women aged 80 and older account for 55% of climate-change-attributable deaths, by the 2010s their per-capita rates were nearly identical to those of men. In other words, the fact that the burden of heat falls primarily on women reflects the fact that women tend to grow older, rather than differing in susceptibility from men. Similar conclusions are reached by Vicedo-Cabrera et al. (2023) and Beck et al. (2024), though both find women slightly more vulnerable than we do.

Why is heat vulnerability declining? The most systematic attempt to answer this is due to Achebak et al. (2023), who tested fourteen candidate drivers in Spain and found an independent association only for air conditioning. This cannot explain the falling vulnerability in the Netherlands, given its limited uptake during our study period (TNO, 2024). Another prominent explanation is general improvements in population health, but the evidence here is contradictory. Huber et al. (2025) identify life expectancy as the strongest predictor of declining vulnerability, while Achebak et al. (2023) find it is unrelated. Similarly, evidence on heat plans is inconsistent (Feldbusch et al., 2025; Urban et al., 2025; Williams et al., 2019). The existing literature thus offers no satisfactory explanation for declining heat vulnerability.

Our study adds two pieces of evidence that can help build a better understanding of declining heat vulnerability. We see a decline that started long before the 2007 Dutch Heat Plan, which argues against the plan as the primary driver. We also see asymmetric trajectories between the sexes, with male rates falling steadily from the start of the series while female rates rose until around 2000 and then fell sharply. This suggests that the drivers do not act on the population as a whole but are sex specific, such as improvements in population health, where men and women followed different paths in smoking and lifestyle (Janssen, 2020).

Additional research is needed to better understand the declining heat vulnerability. First, new candidate drivers must be identified and existing ones tested more rigorously, through richer comparisons across countries that developed differently and across longer periods. Second, moving from all-cause to cause-specific mortality would clarify the pathways through which heat kills and allow interventions to be targeted accordingly. Third, finer spatial and social resolution would reveal the inequalities that national averages hide.

Our estimates align with existing research. Removing the anthropogenic signal cools Dutch warm-season temperatures by about 2.5 ℃ by 2019, matching the KNMI estimate of observed warming since 1901 (van der Wiel et al., 2024). Our aggregate share of 59% sits close to the 53.6% that Huber et al. (2025) report for German cities, and exceeds the 31% implied for the Netherlands by Vicedo-Cabrera et al. (2021). This difference comes down to how the counterfactual time-series is built. Vicedo-Cabrera et al. (2021) relies on climate-model ensembles, which miss recent changes in West-European summer circulation and so underestimate both the warming that occurred and the share attributable to it (Stuart-Smith et al., 2025; Vautard et al., 2023).

As in any empirical work, our conclusions are subject to a number of limitations. First, we use weekly rather than daily data, which smooths out short temperature spikes and likely underestimates both heat deaths and the share attributable to climate change. Second, we use all-cause mortality, so we cannot separate deaths caused by heat, such as cardiovascular and respiratory ones, from those unrelated to it. Third, our counterfactual rests on a scaling factor *α* that translates global warming into Dutch warming, following standard event-attribution protocols (Philip et al., 2020; van Oldenborgh et al., 2021). Our intervals propagate the sampling uncertainty in *α* but not this structural uncertainty, because *α* is estimated against the anthropogenic part of global warming alone and so cannot absorb slow natural swings in climate. Setting *α* to either end of its confidence interval changes the size of the climate-change share but not its sign, its upward trend, or the ordering of age and sex groups. Fourth, our null-mortality check showed that the method becomes unstable when no real heat signal is present, so intervals for strata with weak signals, particularly the 65–79 group, should be read with caution. Fifth, the 80+ group has no upper bound and becomes increasingly female at the oldest ages, so our per-stratum rates do not adjust for the age mix within it. They may therefore understate risk in the oldest women, and comparisons between the sexes at these ages are not age-adjusted.

Human activity is driving global warming toward levels beyond anything societies have experienced. We showed that this warming is already responsible for the majority of heat deaths in the Netherlands. Urgent political action is needed to prevent further loss of life, both by better adapting to existing levels of warming and by mitigating further climate change.

## Acknowledgments

JMBH was supported by the Dutch Research Council (NWO) under VENI grant number 221G.110. AF and RW were funded by the Centre for Urban Mental Health, a research priority area at the University of Amsterdam, and by the REACH Nexus consortium funded by the Dutch Research Council (NWO, project number 24.411).

## Data availability statement

All data and analysis code supporting the findings of this study are openly available in the Open Science Framework repository at https://osf.io/d6rme (DOI: 10.17605/OSF.IO/D6RME).

The repository contains the R scripts for data preprocessing, attribution and robustness analyses, the derived weekly analysis panel, and archived copies of the publicly available source data. The original data are available from Statistics Netherlands, KNMI, NASA GISTEMP, the IGCC project and NOAA’s Physical Sciences Laboratory as described in Appendix A.1.

## Author contributions

**Adam Finnemann:** conceptualization, methodology, software, formal analysis, data curation, visualization, writing — original draft, writing — review and editing. **Fabian Dablander:** conceptualization, methodology, writing — review and editing. **Reinout Wiers:** writing — review and editing. **Jonas Haslbeck:** conceptualization, methodology, writing — review and editing. All authors read and approved the final manuscript.

## Ethics approval

Not required. The study uses only aggregate, publicly available mortality and climate statistics and involves no individual-level data and no human participants.

## A Detailed methods

This appendix gives the full technical specification of the three stages summarized in the Methods. Section A.1 lists the data sources, Section A.2 the remaining details of the epidemiological model, Section A.3 the construction of the counterfactual temperatures, and Section A.4 the attribution step and its uncertainty propagation.

### A.1 Data sources

Weekly all-cause mortality counts stratified by age group and sex were obtained from Statistics Netherlands (CBS; https://opendata.cbs.nl/#/CBS/nl/dataset/70895ned/table). Weekly mean and maximum temperatures were constructed from national station data provided by the Royal Netherlands Meteorological Institute (KNMI). Annual population counts by sex and age group were obtained from CBS (https://www.cbs.nl/en-gb/figures/detail/85510ENG) and used as an offset to account for changing population size.

To compute counterfactual temperatures, we used monthly observed global mean surface temperature (GMST) from NASA GISTEMP (https://data.giss.nasa.gov/gistemp/) and estimates of the global warming component from the Indicators of Global Climate Change (IGCC) project, specifically the GMST time series that splits observed warming into a natural and an anthropogenic component, the latter combining greenhouse gases and other human influences on the climate, with associated 5th–95th percentile uncertainty bounds (Forster et al., 2025). To account for internal climate variability when estimating the Netherlands-to-global scaling factor, we additionally used monthly indices of the El Niño–Southern Oscillation (Niño3.4 SST anomaly) and the North Atlantic Oscillation from NOAA’s Physical Sciences Laboratory (https://psl.noaa.gov/data/climateindices/list/). All data used in this study are freely and publicly available, with archived versions and download scripts provided in our OSF repository (https://osf.io/d6rme); see the data availability statement.

### A.2 Stage 1: model specification details

#### Lag window

We use a two-week lag window because heat elevates mortality both in the week of exposure and in the weeks that follow, while longer windows increasingly capture deaths that would have occurred shortly regardless. Stuart-Smith et al. (2025) use a comparable 10-day window.

#### Flexibility of the time terms

The cyclic cubic spline of week-of-year has flexibility set by *k*_week_ and the smooth of calendar time by *k*_time_. Our main specification uses *k*_week_ = 20 and *k*_time_ = 150. Because the appropriate level of control is not self-evident, we varied both, refitting the model with no within-season spline at all (*k*_time_ = 40), with a minimal seasonal spline (*k*_week_ = 5, *k*_time_ = 40), and with a more flexible trend (*k*_week_ = 20, *k*_time_ = 300). Results are reported in Section B.2.

### A.3 Stage 2: counterfactual temperature details

#### Treatment of the anthropogenic GMST series

We take the 5th, 50th, and 95th percentiles of ΔGMST_ANT_(*y*) from the published IGCC ensemble. The central estimate uses a 4-year centered running mean of the 50th percentile to suppress ENSO-driven interannual noise in the anthropogenic signal, following standard attribution protocol (Philip et al., 2020; van Oldenborgh et al., 2021), while the unsmoothed 5th–95th range is retained as the uncertainty bound.

#### Identification of the scaling factor *α*

Identifying *α* requires separating the Dutch temperature response to global warming from other sources of month-to-month variation. We therefore control for the El Niño–Southern Oscillation (Niño3.4) and the North Atlantic Oscillation, absorb the within-season seasonal cycle with month fixed effects, and cluster standard errors by year to account for within-year serial correlation. In the regression equation given in the Methods, *β*_0_ is the intercept and *γ* and *δ* the coefficients on the Niño3.4 and NAO indices. The constant-*α* assumption is examined in

Section B.3: we test its stationarity over time via a ΔGMST_ANT_ *×* year interaction with cluster-robust inference and rolling-window estimates, and we examine whether *α* varies across the temperature distribution using quantile regression (qgam) across the 5th–95th percentiles.

#### Uncertainty propagation

Uncertainty in the counterfactual temperatures has two sources: the anthropogenic GMST estimate and the scaling factor *α*. We combine them using Monte Carlo simulations. For each year *y*, we take *n*_sim_ = 2,000 draws of ΔGMST_ANT_(*y*) (mean = 4-year smoothed IGCC median, SD from the unsmoothed 5th–95th range) and of *α* (mean and SE from the warm-season regression), multiply them to obtain draws of the Dutch warming shift Δ*T*_NL,ANT_(*y*), and summarize the resulting distribution by its mean and standard deviation. These two numbers per year are passed forward as the climate-uncertainty inputs to the attribution stage. For visualization, we also retain the 5th, 50th, and 95th percentiles of the shift distribution, which define low, central, and high counterfactual weekly temperature series.

### A.4 Stage 3: attribution details

#### Aggregation

Stratum-level annual estimates are aggregated to national annual totals, and further summed across years to obtain cumulative totals for comparison with prior work. Both AN_heat_ and AN_cc_ are reported per stratum as crude rates and combined into age–sex standardized national totals.

#### Joint uncertainty propagation

Our attribution estimates are subject to two sources of uncertainty: (i) the epidemiological exposure–response function, captured by the variance–covariance matrix of the fitted cross-basis coefficients, and (ii) the counterfactual temperature construction, captured by the mean and standard deviation of the Dutch warming shift derived in Stage 2. We propagate both sources jointly via Monte Carlo with *n*_sim_ = 1,000 draws. In each draw we sample a full exposure– response curve for that year from the fitted Stage 1 model by drawing coefficients from a multivariate normal distribution centered on the fitted values and using the model’s estimated coefficient uncertainty, 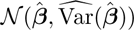) together with a climate shift Δ*T*_NL,ANT_(*y*) *∼ N* (*µ*_Δ*T*_ (*y*)*, σ*_Δ*T*_ (*y*)). We then recompute AN_heat_ under observed temperatures and AN_heat,cf_ under the shifted (counterfactual) temperatures using the *same* coefficient draw, so that AN_cc_ is obtained from a paired difference. Uncertainty intervals for each stratum-year are the 5th and 95th percentiles of the resulting draws, with the 50th percentile reported as the central estimate. National intervals are obtained by summing stratum-specific draws across strata for each year before taking percentiles, ensuring uncertainty propagates coherently through aggregation.

## B Robustness and supporting analyses

Here we test whether our results depend on key modeling choices, and report the supporting analyses referred to in the main text. In Section B.1 we replace weekly mean temperature with weekly maximum temperature. In Section B.2 we vary the seasonal control. In Section B.3 we test whether the scaling factor *α* is constant over time and across the temperature distribution. In Section B.4 we run the method on a series with no heat signal, and in Section B.5 we re-run the attribution across the confidence interval of *α*. Section B.6 reports the climate-change share over time, Section B.7 the pre-attribution screening of every stratum, and Section B.8 heat-mortality rates by sex and age.

### B.1 Sensitivity analysis using weekly maximum temperature (TMAX)

As a sensitivity check we repeated the full attribution analysis using weekly maximum temperature (TMAX) in place of weekly mean temperature, reported on the same rate scale as the main text. Figures S1 and S2 show that the national and stratum-level patterns are qualitatively unchanged: a declining standardized heat burden after the late 1990s, a growing climate-change share, and a burden concentrated at ages 80+ in both sexes.

**Figure S1:**
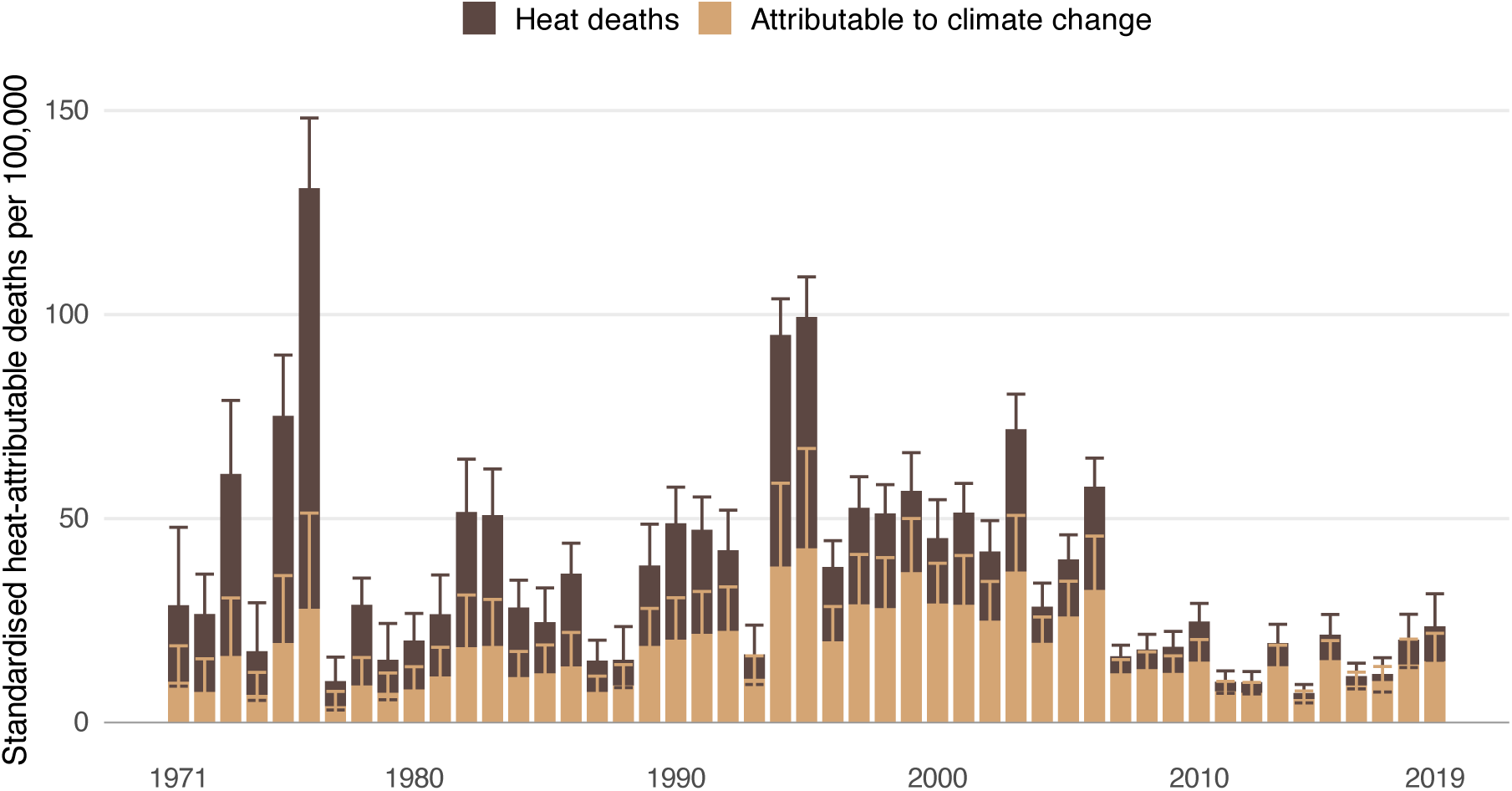
Standardized heat-attributable deaths per 100,000 per warm season in the Netherlands (1971–2019) using weekly maximum temperature (sensitivity analysis), decomposed into deaths attributable to climate change (sand) and remaining heat deaths (dark brown). Rates are age-standardized to the period-average age–sex structure. Error bars show 90% uncertainty intervals.

**Figure S2:**
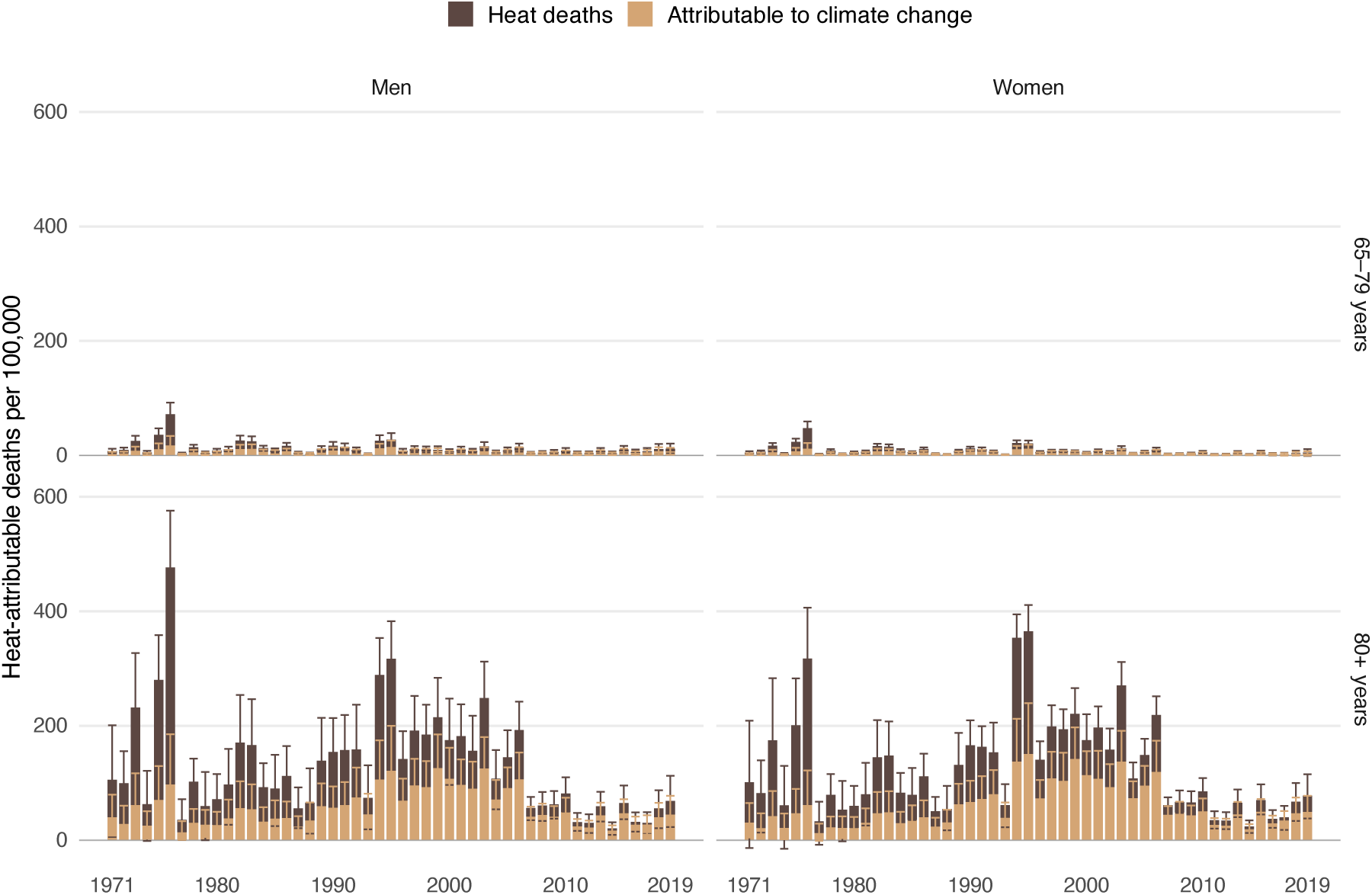
Heat-attributable deaths per 100,000 per warm season by sex and age group (1971–2019) using weekly maximum temperature (sensitivity analysis), decomposed into deaths attributable to climate change (sand) and remaining heat deaths (dark brown). Rates are per 100,000 of each stratum’s own population. Error bars show 90% uncertainty intervals. Only strata with a robust temperature– mortality association are shown.

### B.2 Sensitivity to seasonal control specification

Because the warm season lacks the strong non-temperature seasonal confounders that motivate seasonal adjustment in full-year analyses, the choice of seasonal spline is not self-evident. We refitted the model under four specifications: the original (*k*_week_ = 20, *k*_time_ = 150), a minimal version (*k*_week_ = 5, *k*_time_ = 40), a trend-only version (*k*_time_ = 40, no within-season spline), and a heavy version (*k*_week_ = 20, *k*_time_ = 300). Figure S3 shows that the absolute burden is lower under the less flexible specifications, by up to a third in 2010–2019. The climate-change share, however, is stable across all four, and the main results of a rising climate-change share and a declining burden after the late 1990s remain unchanged.

**Figure S3:**
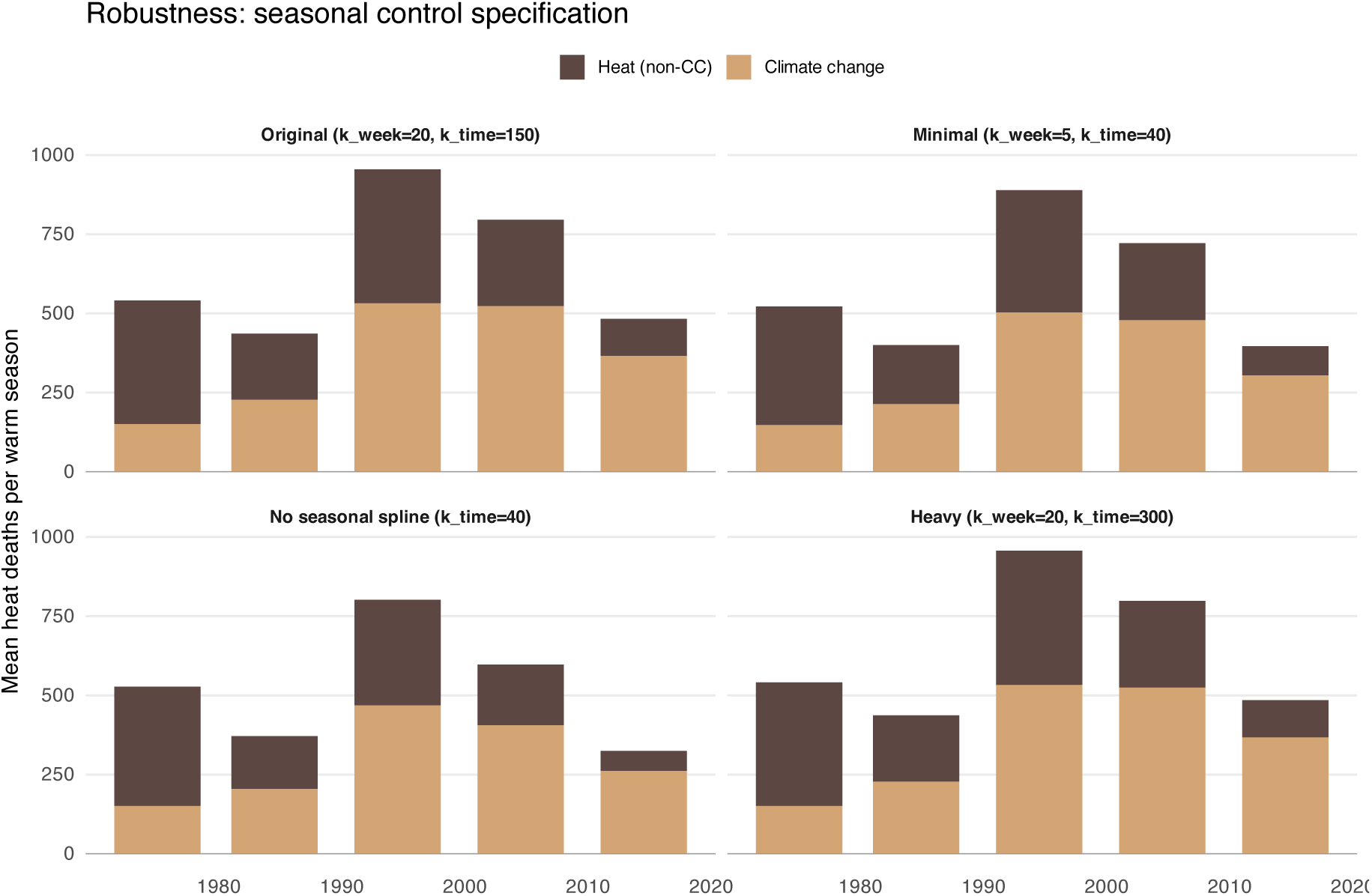
Sensitivity of heat-attributable and climate-change-attributable deaths to the seasonal control specification. Each panel shows period-mean heat deaths per warm season decomposed into the climate-change share and the remainder.

### B.3 Scaling factor ***α***: stationarity and distributional robustness

The attribution pipeline assumes a constant Netherlands-to-global warming scaling factor *α*. We tested this assumption in two ways: (i) a stationarity check by adding a ΔGMST_ANT_ *×* year interaction to the warm-season *α* regression with cluster-robust standard errors, complemented by rolling-window estimates across the study period (Figure S4), and (ii) a distributional check using quantile regression (qgam) to estimate *α* across the 5th–95th percentiles of the Dutch warm-season temperature distribution (Figure S5). Both use the same anthropogenic warming regressor as the main analysis, so the parameter tested is the one the counterfactual applies. Neither test indicated meaningful departures from a constant *α*: the interaction term was not significant (*p* = 0.56), rolling-window estimates remained within their 95% confidence bands, and quantile-specific estimates were stable across the distribution, apart from a slight rise at the uppermost quantiles that stays well within the confidence band of the pooled estimate. Were that rise real, a single warm-season *α* would under-cool the counterfactual at the hottest weeks, making the attributed share conservative. These checks support using a single warm-season *α* in the main analysis.

**Figure S4:**
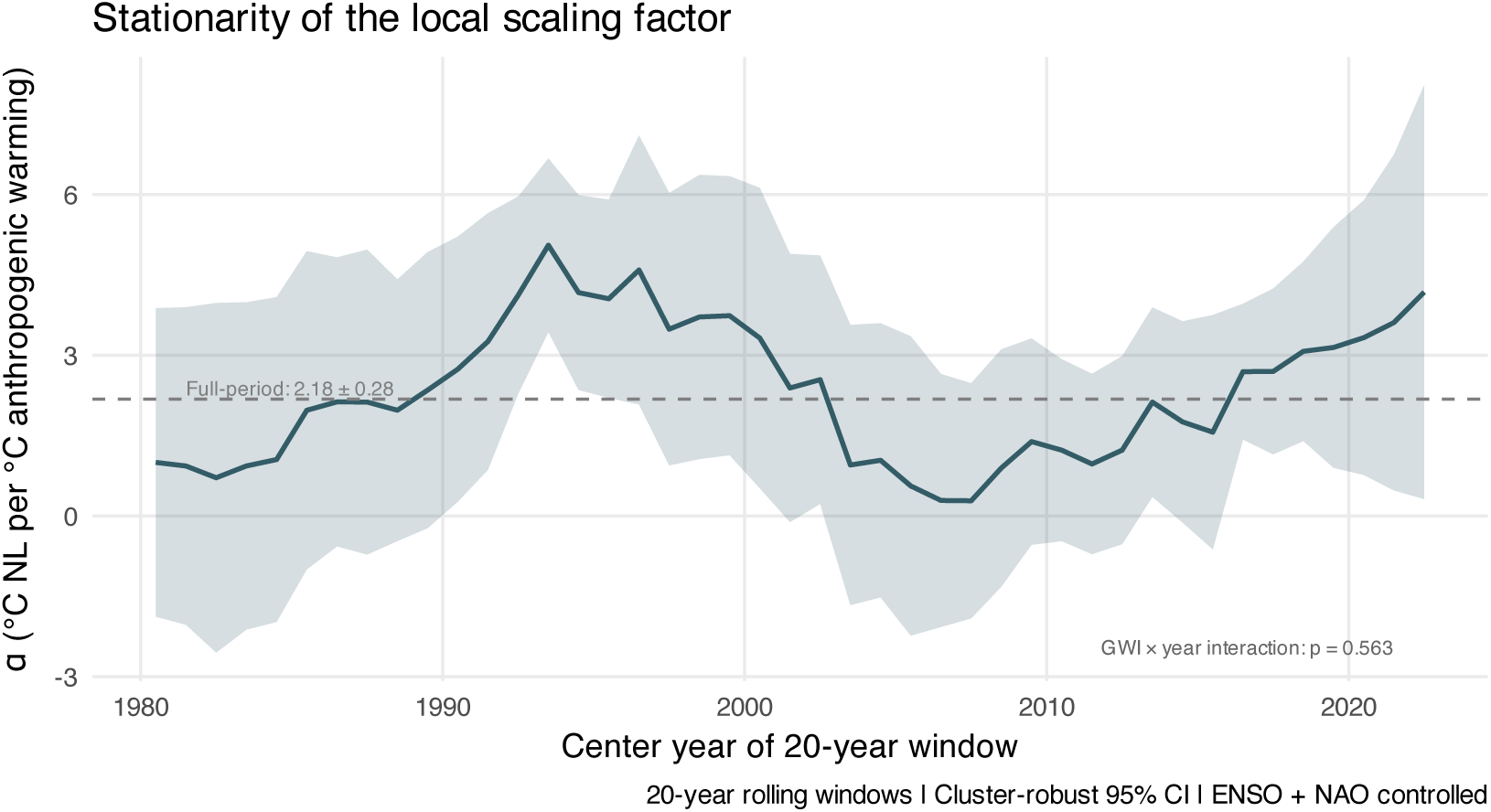
Stationarity of the Netherlands-to-global scaling factor *α*. Rolling-window estimates of *α* with cluster-robust 95% confidence intervals (ENSO and NAO controlled). The dashed line shows the full-period estimate; annotation reports the *p*-value of the ΔGMST_ANT_ *×* year interaction.

**Figure S5:**
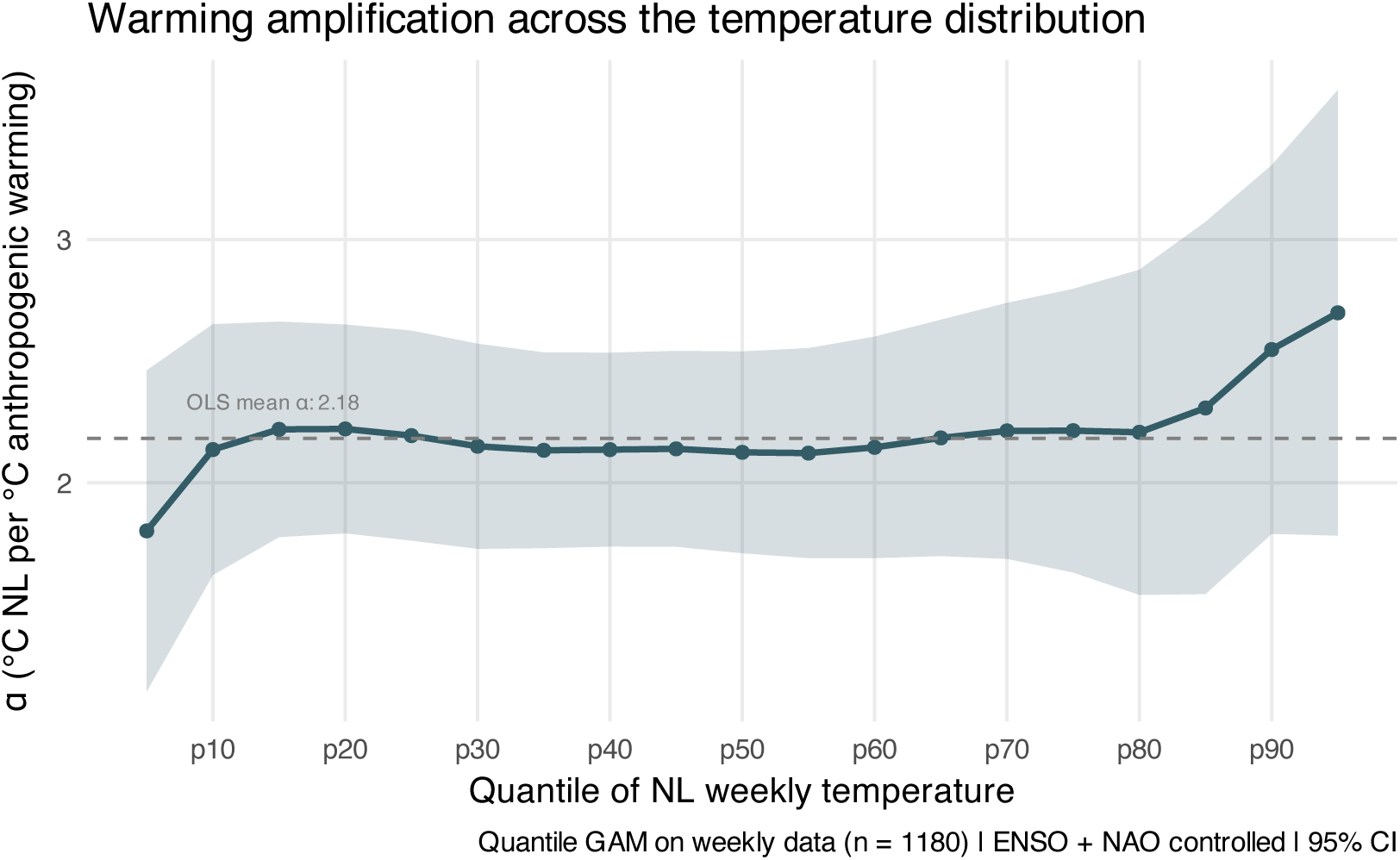
Warming amplification across the Dutch warm-season temperature distribution. Quantile-specific *α* estimated by quantile GAM (qgam) on weekly data at grid quantiles from the 5th to the 95th percentile, with 95% confidence bands. The dashed line shows the OLS mean *α*.

### B.4 Null-mortality robustness

To verify that our pipeline does not manufacture a heat signal from temperature alone, we re-ran the full workflow on a synthetic series in which weekly deaths were replaced by Poisson draws around the pooled warm-season mean, independent of temperature. Figure S6 shows that the year-specific MMT snaps between discrete boundary values of the temperature distribution rather than fluctuating around a stable interior minimum. This reflects the fact that MMT is only well defined when the RR curve is V-shaped; under the null the fitted cross-basis is near-linear with slowly drifting slope, so its minimum is attained at the boundary of the search range and flips abruptly when the slope changes sign. The same pathology appears in the real 0–64 strata, supporting their exclusion.

**Figure S6:**
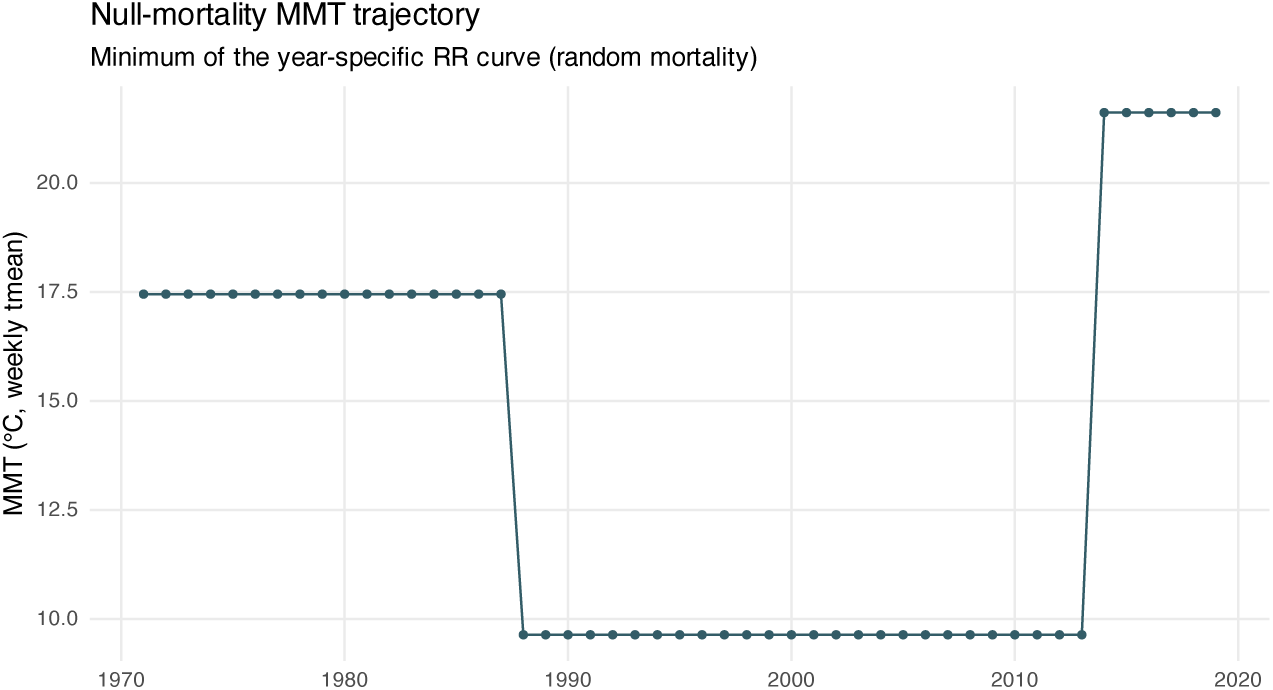
Year-specific MMT under the null-mortality robustness run (weekly deaths replaced by Poisson draws around the pooled mean). MMT snaps between boundary values rather than fluctuating around a stable interior minimum, reflecting near-linear RR curves in the absence of a true temperature–mortality link.

### B.5 Sensitivity of attribution to the magnitude of ***α***

The checks above test whether *α* is stable. Here we ask how much the attribution results depend on its value. Because the counterfactual cooling is Δ*T*_NL,ANT_(*y*) = *α ·* ΔGMST_ANT_(*y*), it is linear in *α*, so an alternative *α* simply rescales the per-year shift. We re-ran the attribution under four values: the central warm-season estimate (*α* = 2.19, reproducing the main results), its lower and upper 95% confidence bounds (1.65 and 2.74), and a value obtained by regressing monthly Dutch temperature on *total* observed GMST rather than on its anthropogenic component, with the same ENSO, NAO, and month-fixed-effect controls (*α* = 2.03). The last addresses the concern that the choice of regressor drives the result. Regressing on total GMST returns a slightly smaller *α* and lowers the cumulative climate-change share by 2.7 percentage points, from 59.1% to 56.4%. Table S1 summarizes the results. Total heat-attributable deaths are unaffected by *α* (cumulative 31,551). The climate-change share scales with *α* in magnitude but, in every scenario, stays positive, rises across the three periods, and preserves the age–sex ordering (women aged 80+ *≈* 55%, men aged 80+ *≈* 24% of climate-change-attributable deaths).

**Table S1:** Climate-change attribution under alternative values of the Netherlands-to-global scaling factor *α*. Cumulative heat-attributable deaths are fixed at 31,551 across all scenarios, since *α* affects only the counterfactual. “CC share” is the cumulative climate-change share of heat-attributable deaths; the period columns give the same share within each period; the final two columns give each stratum’s share of all climate-change-attributable deaths.

| Scenario | $\alpha$ | Climate-change share (%) | | | | Share of CC deaths (%) | |
| --- | --- | --- | --- | --- | --- | --- | --- |
|  |  | Overall | 1971–94 | 1995–09 | 2010–19 | Women 80+ | Men 80+ |
| $\alpha$ lower 95% CI | 1.65 | 48.9 | 36.8 | 54.9 | 67.4 | 54.8 | 24.4 |
| $\alpha$ central (main) | 2.19 | 59.1 | 45.9 | 66.1 | 78.5 | 54.8 | 24.3 |
| $\alpha$ upper 95% CI | 2.74 | 67.4 | 53.8 | 74.8 | 86.4 | 54.8 | 24.3 |
| $\alpha$ vs. total GMST | 2.03 | 56.4 | 43.4 | 63.0 | 75.6 | 54.8 | 24.3 |

### B.6 Climate-change share over time

The share of heat deaths attributable to climate change rose steadily across the study period, from about one-third in the early 1970s to roughly three-quarters by the 2010s (Figure S7).

**Figure S7:**
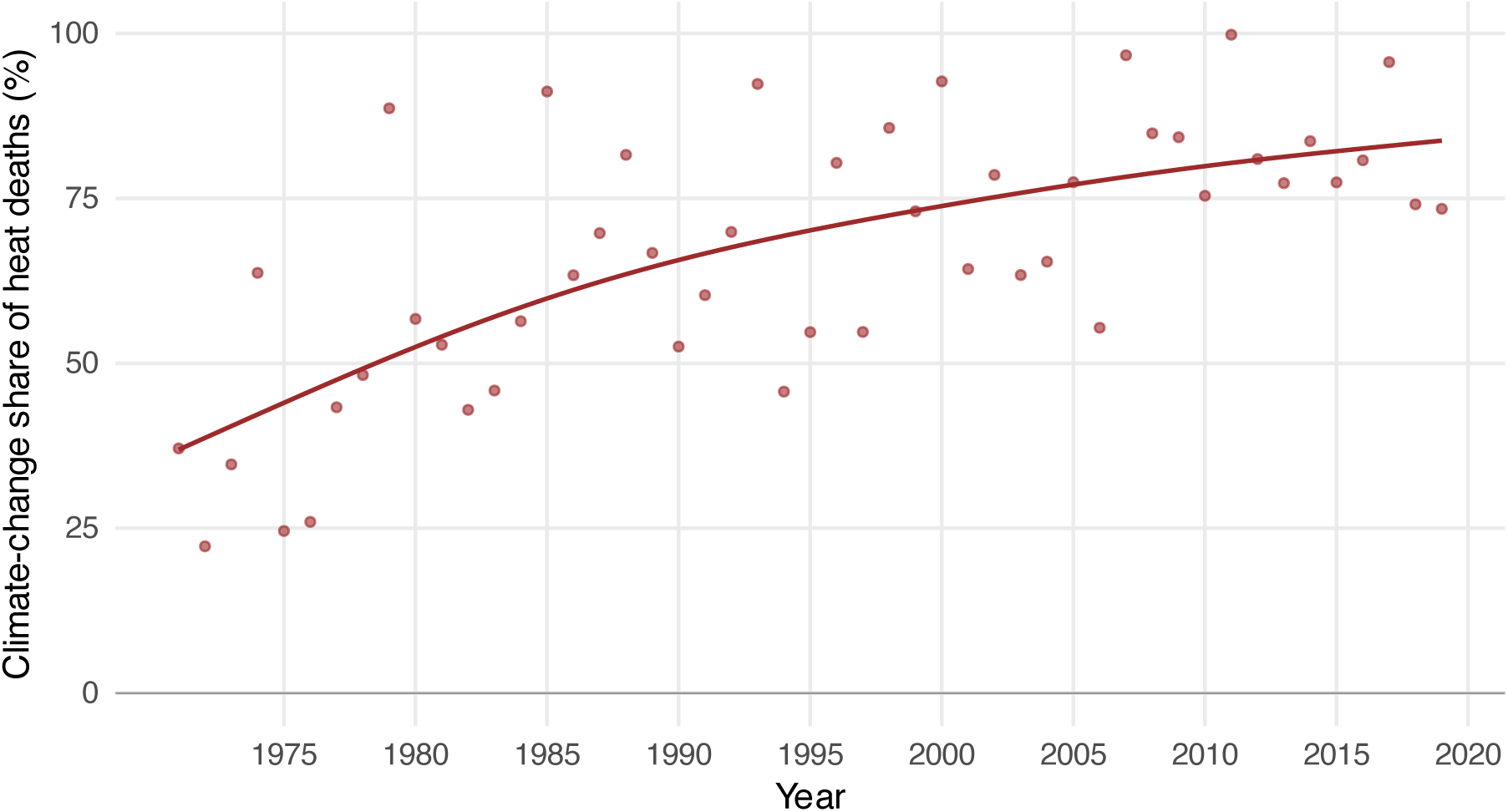
Annual climate-change share of warm-season heat deaths (%), 1971–2019, with a smoothed trend (penalized spline). The share rises steadily across the study period, from around one-third in the early 1970s to roughly three-quarters by the 2010s, an average increase of about 9 percentage points per decade.

### B.7 Pre-attribution screening: RR curves and MMT stability

Because our method is only well-defined when there is a clear heat–mortality association, we screened every stratum for V-shaped exposure–response curves and stable, interior MMT estimates before running the attribution analysis. Figure S8 shows the fitted RR curves and Figure S9 the year-specific MMT trajectories for all strata. The 65–79 and 80+ strata exhibit V-shaped curves with stable interior MMTs around 16–19 ℃, whereas the 0–64 strata show near-linear RR curves and unstable, boundary-snapping MMTs consistent with a weak or absent heat–mortality association, and were therefore excluded from the main attribution analysis.

**Figure S8:**
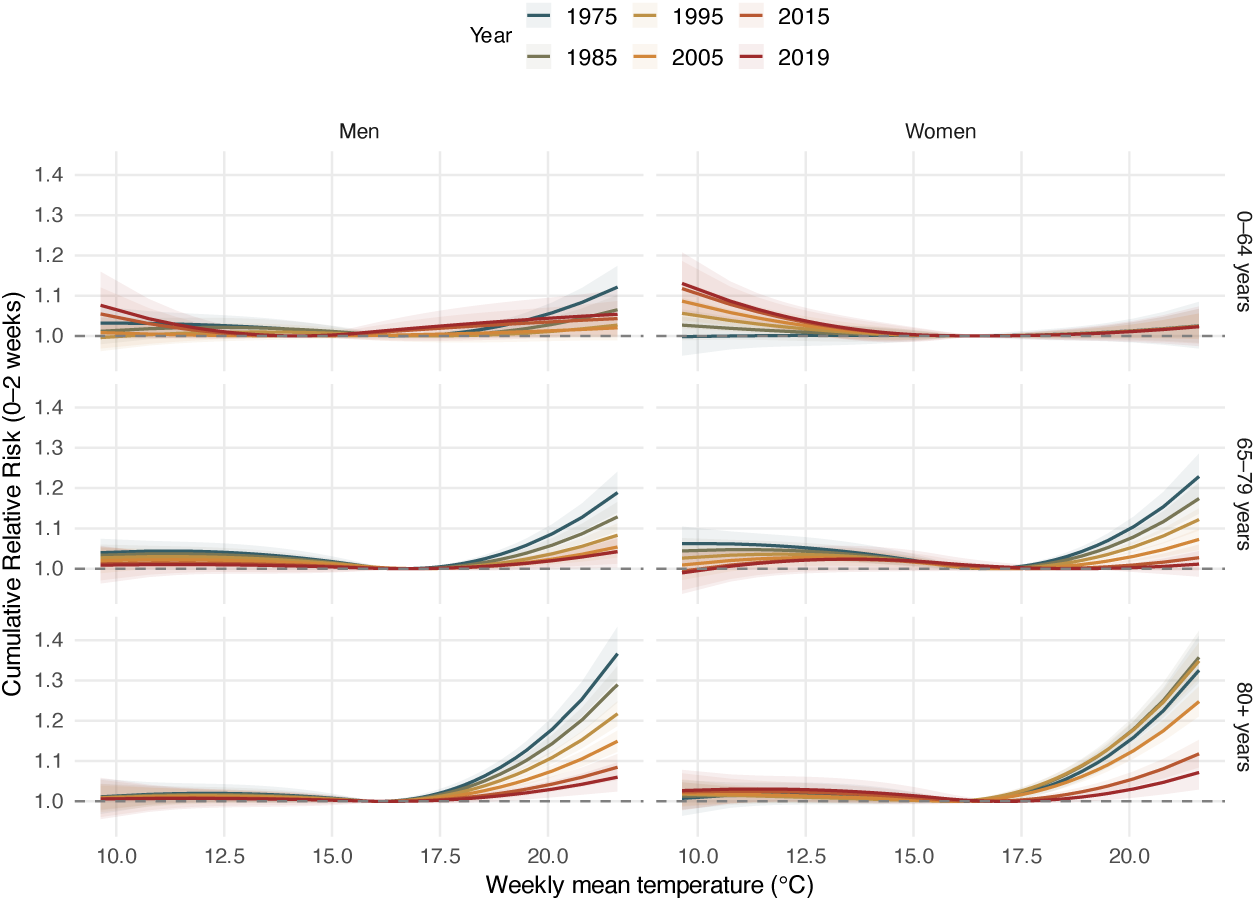
Time-varying temperature–mortality RR curves (TMEAN) for all sex *×* age strata. Curves show year-specific cumulative RR over lags 0–2 weeks (colored by year), centered at the year-specific MMT, with 95% CIs. The 0–64 strata display near-linear rather than V-shaped curves.

**Figure S9:**
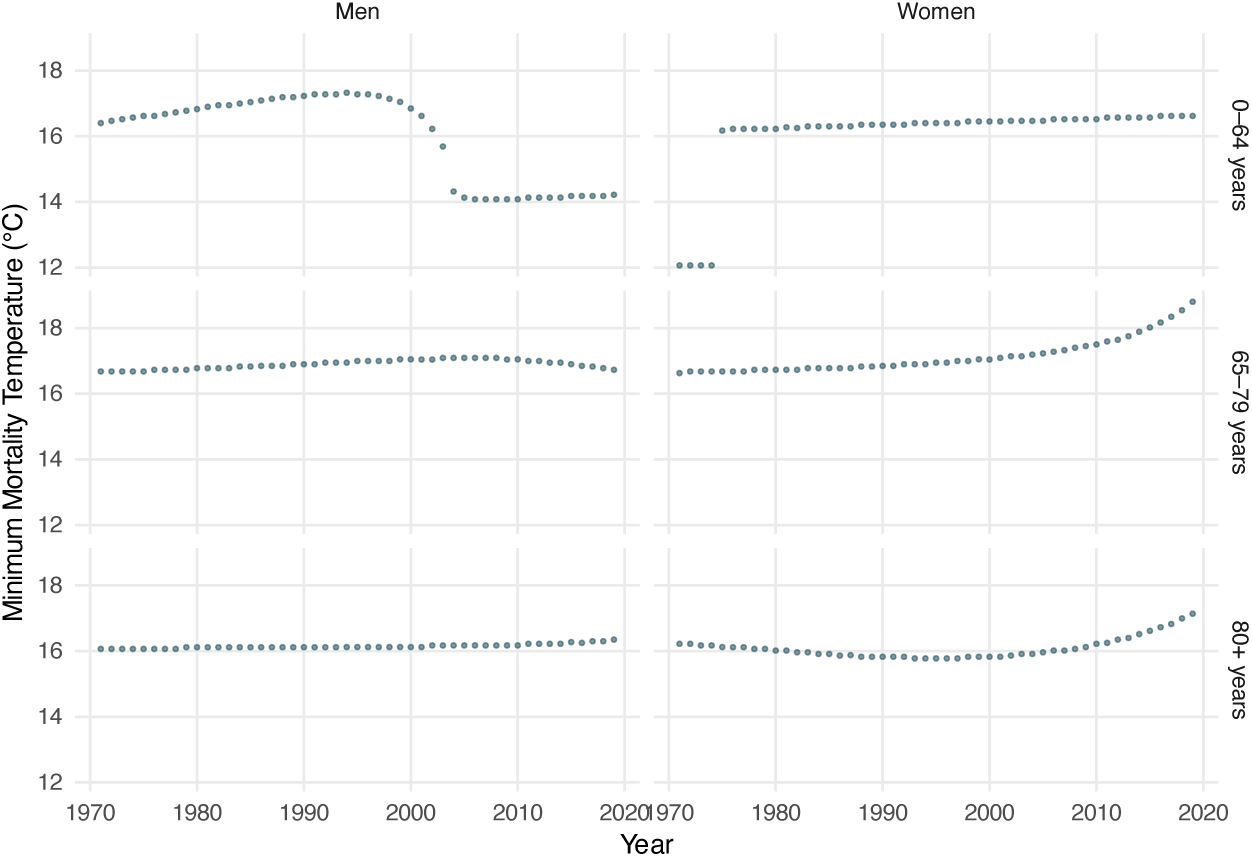
Year-specific minimum mortality temperature (MMT) for all sex *×* age strata (TMEAN). The 65+ strata show stable interior MMTs around 16–19 ℃; the 0–64 strata show unstable, drifting or boundary-snapping MMTs consistent with an ill-defined minimum.

### B.8 Heat-mortality rates by sex and age over time

**Table S2:** Heat-attributable mortality rate per 100,000 by sex and age group, selected years. Rates are derived from smoothed annual estimates under observed temperatures. W/M is the ratio of the female to the male rate, with values above one indicating higher female risk.

| Year | Aged 80+ |  |  | Aged 65–79 |  |  |
| --- | --- | --- | --- | --- | --- | --- |
|  | Men | Women | W/M | Men | Women | W/M |
| 1975 | 145 | 97 | 0.67 | 20.1 | 14.4 | 0.72 |
| 1985 | 125 | 115 | 0.92 | 15.8 | 10.3 | 0.65 |
| 1995 | 109 | 148 | 1.37 | 11.4 | 7.2 | 0.63 |
| 2000 | 104 | 150 | 1.44 | 9.6 | 6.0 | 0.62 |
| 2005 | 88 | 120 | 1.37 | 8.2 | 5.0 | 0.61 |
| 2010 | 74 | 86 | 1.17 | 6.5 | 3.8 | 0.58 |
| 2015 | 61 | 59 | 0.97 | 4.7 | 2.5 | 0.53 |
| 2019 | 53 | 44 | 0.82 | 3.8 | 1.7 | 0.45 |

